# Dynamics of *Plasmodium falciparum* asymptomatic asexual and sexual stages across transmission seasons in a rural and high-transmission setting in Burkina Faso: a two-year longitudinal cohort study with cross-sectional surveys

**DOI:** 10.64898/2026.01.09.26343521

**Authors:** D. Florence Ouédraogo, Vera Kühne, Rouamba Toussaint, Pieter Guetens, Aida Millogo, Ana Moreno-Murillo, Yasmina Drissi-El Boukili, Ana Chopo-Pizarro, Neus Ràfols, Erin Sauve, Adina Asim, Abdoul Rahim Ouedraogo, Abdoulaye Ouédraogo, Karim Derra, M. Athanase Somé, Esther Hien, Aly Savadogo, Hermann Sorgho, Halidou Tinto, Anna Rosanas-Urgell, H. Magloire Natama

**Affiliations:** Unité de Recherche Clinique de Nanoro, Institut de Recherche en Sciences de la Santé, Direction régionale du Nando, Nanoro, Burkina Faso; Department of Biomedical Sciences, Institute of Tropical Medicine, Antwerp, Belgium; Laboratoire de Biochimie et d’Immunologie Appliquées (LaBIA), Ecole Doctorale Sciences et Technologies, Université Joseph Ki-Zerbo, Ouagadougou, Burkina Faso

## Abstract

**Background:** Malaria caused by *Plasmodium falciparum* remains a major global health challenge. Although clinical disease results from asexual blood-stage replication, transmission depends on gametocyte carriage. We characterized the seasonal dynamics of the human infectious reservoir in a seasonal, high-transmission setting in Burkina Faso.

**Methods:** In this longitudinal cohort study with cross-sectional surveys, conducted between March, 2019, and March, 2021, we followed 864 individuals of all ages from four villages in the Nanoro health district, Burkina Faso, selected from the health and demographic surveillance system by age group; pregnant women and individuals with underlying disease were excluded. Participants were actively screened for asymptomatic infection three times per year and passively monitored for clinical malaria. *P. falciparum* infections were detected by light microscopy and quantitative PCR targeting parasite DNA and gametocyte-specific RNA. Primary outcomes were the prevalence of asymptomatic infection and gametocyte carriage across transmission seasons.

**Findings:** Clinical and asymptomatic infections both showed marked seasonality, peaking in the high-transmission season (HTS, July–December): 685/881 (77·8%) to 748/814 (91·8%) of clinical cases occurred in this period. Asymptomatic infections were prevalent during both the low-transmission season (LTS; 262/663 [39·5%] to 379/811 [46·7%]) and HTS (324/596 [54·4%] to 345/544 [61·0%]), with a higher proportion of gametocyte carriage during LTS (147/261 [55·1%] to 223/367 [60·8%]); compared to HTS (103/298 [34·7%] to 174/314 [55·1%]). Age was the strongest predictor of both asymptomatic infection and gametocyte carriage; children aged 5–12 years formed the largest, persistent reservoir across seasons. Transmission potential was highest in children younger than 13 years and peaked at during LTS.

**Interpretation:** School-aged children represent a key, under-targeted reservoir sustaining malaria transmission across seasons in this high-transmission setting. Targeted interventions delivered during the low-transmission season, alongside existing control measures, could reduce gametocyte carriage and shrink the infectious reservoir before the onset of the HTS.

**Funding:** Belgium Directorate General for Development Cooperation (DGD); Research Foundation Flanders (FWO).

## INTRODUCTION

Transmission of the protozoan parasite, *Plasmodium falciparum* from an infected human host to a female *Anopheles* mosquito depends on the presence of sexual parasite stages (gametocytes) in the host’s blood circulation. Circulating gametocyte densities reflect prior asexual parasite dynamics but also vary substantially between individuals and over the course of infection. This variability is influenced by multiple factors such as antimalarial treatment and hemoglobin beta genotype.^1,2^

Characterising the human infectious reservoir in specific malaria-endemic populations is crucial for context-specific malaria control interventions. For targeted transmission-reducing interventions it is essential to know whether specific demographics are more infectious, if there are foci associated with higher infectiousness and if infectiousness follows a specific temporal pattern. Moreover, it is important to understand where current local malaria control strategies effectively target the identified reservoir.

Burkina Faso, with a high and seasonal malaria transmission, is within the top ten most contributing countries to the global malaria burden with 8·3 million cases recorded in health facilities in 2024 and 2.7% of global malaria-related deaths.^3^ Current control strategies (mostly tailored to reduce clinical episodes) include: (i) preventive strategies for targeted populations (*e.g*., seasonal malaria chemoprevention (SMC) for children under five years, intermittent preventive treatment during pregnancy with sulfadoxine-pyrimethamine (IPTp-SP) and malaria vaccines for children starting at 5 months since 2024); (ii) the test and treat strategy for case management using artemisinin-based combination therapies (ACTs); and (iii) vector control measures (*e.g*., distribution of long lasting insecticide treated nets (ITNs) and indoor residual spraying).

Evidence from both longitudinal and cross-sectional studies highlights the role of age in shaping the human infectious reservoir. School-aged children (five–15 years) consistently exhibit the highest prevalence of *P. falciparum* infection and gametocyte carriage and contribute disproportionately to transmission in Burkina Faso and in other endemic countries.^4–6^ Existing SMC in Burkina Faso would miss this age group.

While current “test and treat” strategies detect clinical cases, they miss asymptomatic infections. It is therefore important to understand how missed asymptomatic infections contribute to transmission and whether they remain asymptomatic or progress to disease requiring treatment. A recent study conducted in eastern Uganda, where substantial progress in malaria control has been achieved, showed that the human infectious reservoir consists primarily of asymptomatic microscopically detectable infections, followed by asymptomatic submicroscopic and symptomatic infections.^7^ Across settings, asymptomatic infections vary substantially in duration and gametocyte density, with longer-lasting, higher-density infections —more common in older children and adolescents— disproportionately driving transmission ^8^

This study aimed to characterise the prevalence of *P. falciparum* infection and gametocyte carriage across transmission seasons over two consecutive years in a highly seasonal malaria setting including both clinical and asymptomatic infections. We further assessed the longitudinal dynamics of infection, gametocyte carriage, and clinical malaria episodes, and identified demographic groups with the greatest transmission potential using an infectiousness threshold derived from the association between gametocyte density and mosquito infection success from previous membrane feeding studies.

## METHODS

### Study design

We conducted a two-year village-based longitudinal cohort study with cross-sectional surveys (N=864) in Nanoro health district, a rural area located in the Nando (ex Centre-West) region of Burkina Faso (Figure 1A/B). Malaria transmission in the region is seasonal and hyperendemic, with the high-transmission season (HTS) lasting from July to December, and a low-transmission season (LTS) from January to June. Rainfall is highly seasonal: weak precipitation starts in March and passes the threshold of 50 mm/month in May/June, exceeding the 100 mm/month threshold from July to September, no precipitations occur between November and February (Figure 1C, Supplementary Figure 1).

**Figure 1.**
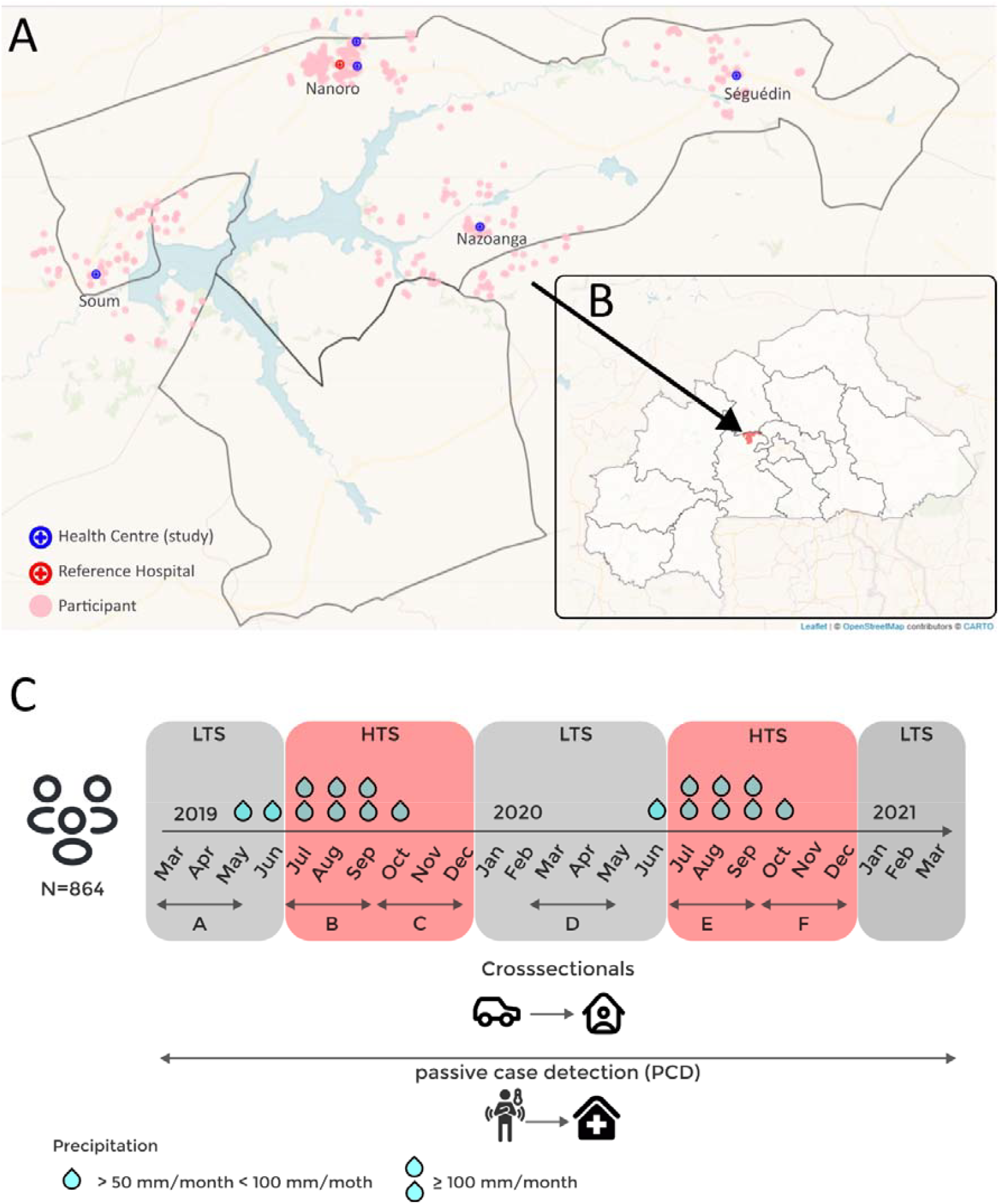
Schematic representation of the study area and design. A) Map of the study area showing location of the four study villages (Soum, Nanoro, Nazoanga and Séguédin), the study health centres (blue cross), the reference hospital (red cross) and the households of the participants (pink dots) within Nanoro Health District (black outlines). B) Location of Nanoro Health District (red) in Burkina Faso (white). C) Schematic representation of the study design with associated precipitation during the low transmission seasons (LTS) (boxes in grey) and the high transmission seasons (HTS) boxes in red. Level of precipitation is shown per month (> 50 mm/month and <100, ≥ 100 mm/month).

The study was conducted from mid-March 2019 to mid-March 2021 with selected participants followed using active case detection (ACD) of asymptomatic infections and passive case detection (PCD) of clinical episodes throughout the study. ACD was implemented through cross-sectional surveys (CSS) conducted by field teams in participant’s homes once during LTS and twice during HTS over two consecutive years (LTS: March-April-May, CSS A&D, HTS: July-August-September, CSS B&E and October-November-December, CSS C&F) (Figure 1C). For PCD, study participants were sensitized at enrolment and during all CSS to visit peripheral health facilities for any illness during the two-year study period. All suspected malaria cases were screened by study nurses appointed to the health centres in the study villages and treated according to national guidelines.

The study was approved by the national ethics committee in Burkina Faso (CERS N°2018-10-131), the institutional review boards of the Institute of Tropical Medicine in Belgium (IRB/AB/AC/027/1261/18), and the University Hospital in Antwerp (UZA) in Belgium (19/06/064). All experiments were performed in accordance with the approved study protocol.

### Participants

Study participants from Nanoro, Nazoanga, Séguédin and Soum villages (Figure 1 A) were selected from the existing health and demographic surveillance system (HDSS) database following methods previously described.^9,10^ Children one to four years old (children younger than five years) were randomly selected according to a computer-generated list. For each child, another paired participant from the same household (HH) was considered for enrolment: either an individual aged five-19 years or >20 years. To minimize sampling bias, two attempts were made to establish contact with a selected child’s family before selecting another child from the list.

Eligibility criteria included residence in selected villages within the HDSS catchment area and willingness to provide written informed consent (or consent by parent(s)/guardian(s) for minors between one and 18 years old). Exclusion criteria included: unwillingness to participate or provide a sample, severe illness requiring hospitalization at the time of recruitment, presence of any underlying disease (*e.g*., cardiac, renal, hepatic), permanent disability that prevents or impedes study participation and/or comprehension, plans to leave the study area, or pregnancy of less than three months.

Written informed consent was obtained from the selected child’s parent/guardian and from HH members willing to be enrolled.

### Procedures

At enrolment, demographic data were collected. Sex assigned at birth was self-reported, with options of female or male. For ACD, a clinical examination (including temperature) was performed during each CSS and additional information on medical history, previous malaria treatment, and ITN physical condition and use was collected by study nurses. Finger prick blood samples collected in EDTA-microtainers, were used to prepare slides for light microscopy (LM), stored for DNA extraction and immediately preserved in RNAprotect Cell Reagent (Qiagen, cat. no. 76526) for RNA extraction. Symptomatic individuals with a positive rapid diagnostic test (RDT) were treated according to national guidelines. If the RDT results were negative, individuals were referred to health facilities. For PCD, participants visiting village health centres underwent the same clinical examination and blood collection procedures.

All slides were examined for *Plasmodium* parasites by LM according to two microscopists with a 3rd read to resolve discrepancies.

*P. falciparum* infection was detected on extracted DNA by targeting *varATS* genes using qPCR, as previously described.^11^ A selection of 10% of samples was shipped to ITM for quality control, demonstrating <10% disagreement in *varATS* qPCR results.

Samples that were *varATS* qPCR positive (CT < 37·2) were subjected to RNA extraction and first strand cDNA synthesis, as previously described.^12,13^ *Pfs25* copies per µl of blood were calculated using a standard curve generated from serially diluted synthetic cRNA controls, as previously described.^14^

Haemoglobin beta sequencing was performed following the PCR conditions adapted from Wittwer *et al*. (2003) ^15^.

All primers used for qPCR and RT-qPCR analysis are listed in Supplementary Table 1.

Clinical malaria (symptomatic infection) was defined as the presence of *P. falciparum* of any density detected by *varATS* qPCR, with axillary temperature ≥37·5°C or history of fever within the previous 24 hours (h). Asymptomatic infection was defined as the presence of *P. falciparum* by *varATS* qPCR without fever in the previous 24 h during ACD. Sub-microscopic infection was defined as infection detected by qPCR but not by LM. Gametocyte carriage was defined as detection of gametocyte stage parasites by RT-qPCR targeting *pfs25* and *pfmget* in *varATS* qPCR positive samples. Sub-microscopic gametocyte carriage was defined as RT-qPCR positivity without gametocytes detected in LM.

### Statistical analysis

Sample size was calculated assuming 30% (LM) and 50% (qPCR) baseline infection prevalence and 50% gametocyte positivity among infections, requiring 750 individuals to detect a 17% difference in gametocyte carriage by HBB genotype (MAF=5%, 80% power); adjusting for 15% anticipated loss-to-follow-up^10^ gave a target of 863 individuals. To avoid bias in population selection in different villages, the proportion of randomized individuals per village was adjusted to achieve a similar proportion of HH involvement, which was estimated at 25% in each village.

All data were processed and stored using software specified in Supplementary Table 2. Entries with missing data relevant to a given analysis were excluded. Descriptive statistics were used to summarize baseline data. Categorical variables were compared using Pearson’s χ^2^-test; continuous variables (medians, IQR) using Kruskal-Wallis with Dunn’s pairwise comparison (Holm-adjusted) and Spearman correlation.

This study employed a mixed-effects logistic regression model approach (*i*.*e*., generalized linear mixed models (GLMMs)) to investigate the prevalence ratio of the factors associated with asymptomatic infections and gametocyte carriage during the CSS. The fixed effects included the following environmental and host parameters: village, ITN use, HBB genotype, sex, and age. CSS and ID were modelled as random effects.

Transmission potential was estimated using mosquito membrane feeding assay data from Ouédrago *et al* 2016 ^17^, collected in Burkina Faso 50-100 km from the study area. Data was stratified to align with the mosquito membrane feeding assay data by season (start wet season = four months from the month with first rainfall (mm/month > one) in season, end wet season = following four months, dry season = remaining four months) and age group (under five, 5-14, 15-30 and >30 years old). The minimum gametocyte/µl value to infect >20 % of mosquitoes was set as the transmission threshold.

Geospatial data were analysed using GPS coordinates of participants’ residence and health centres. Waterbody information was retrieved from https://diva-gis.org/data.html.

In all analyses, *P* values <0·05 were considered statistically significant.

## RESULTS

Of 864 participants enrolled, the majority were children younger than five years (428 [49·1%]) or adults aged ≥20 years (215 [24·7%]); the remainder (221 [25·4%]) were school-aged children or adolescents (Table 1). Most participants owned an insecticide-treated net (632 [72·6%]), though ownership and net condition varied by age and village (Table 1, Figure 2).

**Table 1:** Baseline characteristics of the study participants·.

|  | Overall | 1 – 4 Years | 5 – 12 Years | 13 – 19 Years | ≥ 20 Years | *P*<sup>1</sup> |
| --- | --- | --- | --- | --- | --- | --- |
| N | 864 | 428 | 175 | 46 | 215 |  |
| Sex [number of female (%)] | 498 (57.2) | 205 (47.9) | 75 (42.9) | 23 (50.0) | 195 (90.7) | <0.001 |
| Age [years, median (IQR)] | 5.06 (15.27) | 3.38 (1.61) | 7.21 (2.64) | 15.51 (3.10) | 35.32 (12.11) | <0.001 |
| Participants per village [n(%)] |  |  |  |  |  | 0.3 |
| Nanoro | 376 (43.2) | 188 (43.9) | 66 (37.7) | 27 (58.7) | 95 (44.2) |  |
| Nazoanga | 188 (21.6) | 91 (21.3) | 45 (25.7) | 4 (8.7) | 48 (22.3) |  |
| Séguédin | 134 (15.4) | 66 (15.4) | 31 (17.7) | 5 (10.9) | 32 (14.9) |  |
| Soum | 166 (19.1) | 83 (19.4) | 33 (18.9) | 10 (21.7) | 40 (18.6) |  |
| ITN ownership [n(%)] | 632 (72.6) | 321 (75.0) | 122 (69.7) | 25 (54.3) | 164 (76.3) | 0.011 |
| ITN condition [n(%)] |  |  |  |  |  | >0.9 |
| Good condition<sup>A</sup> | 346 (54.3) | 177 (54.6) | 66 (53.7) | 15 (53.6) | 88 (54.3) |  |
| Torn | 287 (45.1) | 145 (44.8) | 56 (45.5) | 13 (46.4) | 73 (45.1) |  |
| Other | 4 (0.6) | 2 (0.6) | 1 (0.8) | 0 (0.0) | 1 (0.6) |  |
| ITN use<sup>B</sup> [n(%)] | 455 (52.2) | 234 (54.7) | 85 (48.6) | 13 (28.3) | 123 (57.2) | 0.002 |
| Haemoglobin genotype [n(%)] |  |  |  |  |  |  |
| HbAA | 617 (70.8) | 299 (69.9) | 126 (72.0) | 35 (76.1) | 152 (70.7) | >0.9 |
| HbAC | 174 (20.0) | 90 (21.0) | 34 (19.4) | 10 (21.7) | 39 (18.1) |  |
| HbAS | 64 (7.3) | 31 (7.2) | 12 (6.9) | 1 (2.2) | 19 (8.8) |  |
| HbCC | 2 (0.2) | 2 (0.5) | 0 (0.0) | 0 (0.0) | 0 (0.0) |  |
| HbSC | 7 (0.8) | 4 (0.9) | 1 (0.6) | 0 (0.0) | 2 (0.9) |  |

ITN; Insecticide Treated bed Net, <sup>A</sup>ITN good condition: ITN without any physical damage; <sup>B</sup>ITN use: participants who slept under an ITN the night before their enrolment; <sup>1</sup> $\chi^2$ test; Kruskal-Wallis test

**Figure 2.**
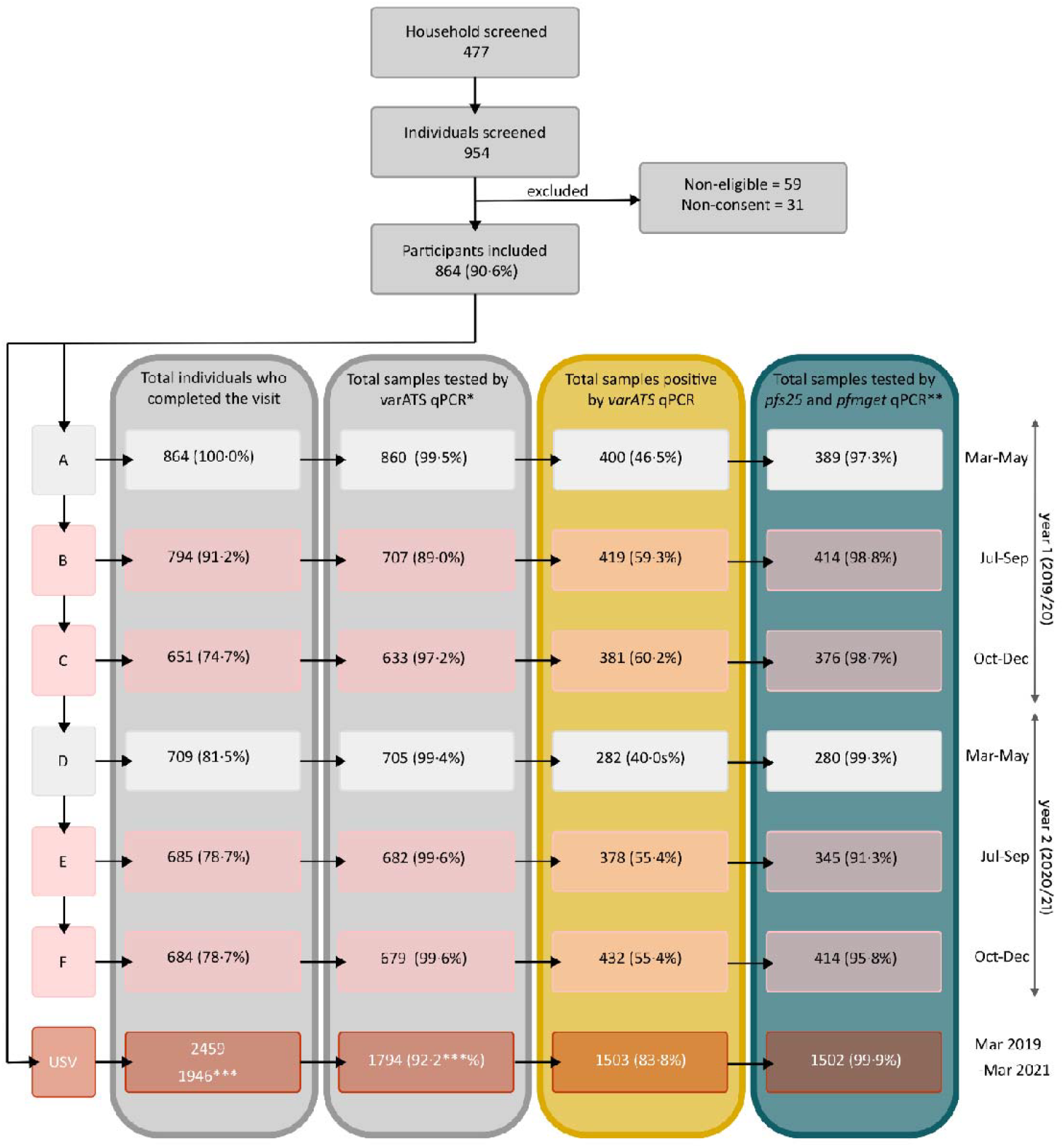
Flow diagram of participant screening, inclusion and follow-up. HTS months highlighted in red-lined boxes and LTS months in grey, Letters A-F represent CSS. *qPCR for the detection of *P. falciparum*; ** qPCR for the detection of *P. falciparum* sexual stages, only performed on *varATS* qPCR positive samples *** with fever or history of fever (subsequent analyses only performed on those), USV, unscheduled visit, HTS, high-transmission season, LTS, low-transmission season, CSS, cross-sectional survey.

Transmission was highly seasonal, with most clinical cases (748/814 [78%] in 2019, 685/881 [91%] in 2020) occurring during HTS (July-December) detected by PCD (Figure 3A). 1497 of 1717 [87·2%] clinical cases were detected by both LM and qPCR (year 1= 686/814 [84·3%], year 2= 811/903 [89·8%]). Peaks in clinical cases occurred earlier in 2019 (September) than in 2020 (October), consistent with earlier onset of rainfall in 2019 (Figure 3A, Supplementary Figure 1).

**Figure 3.**
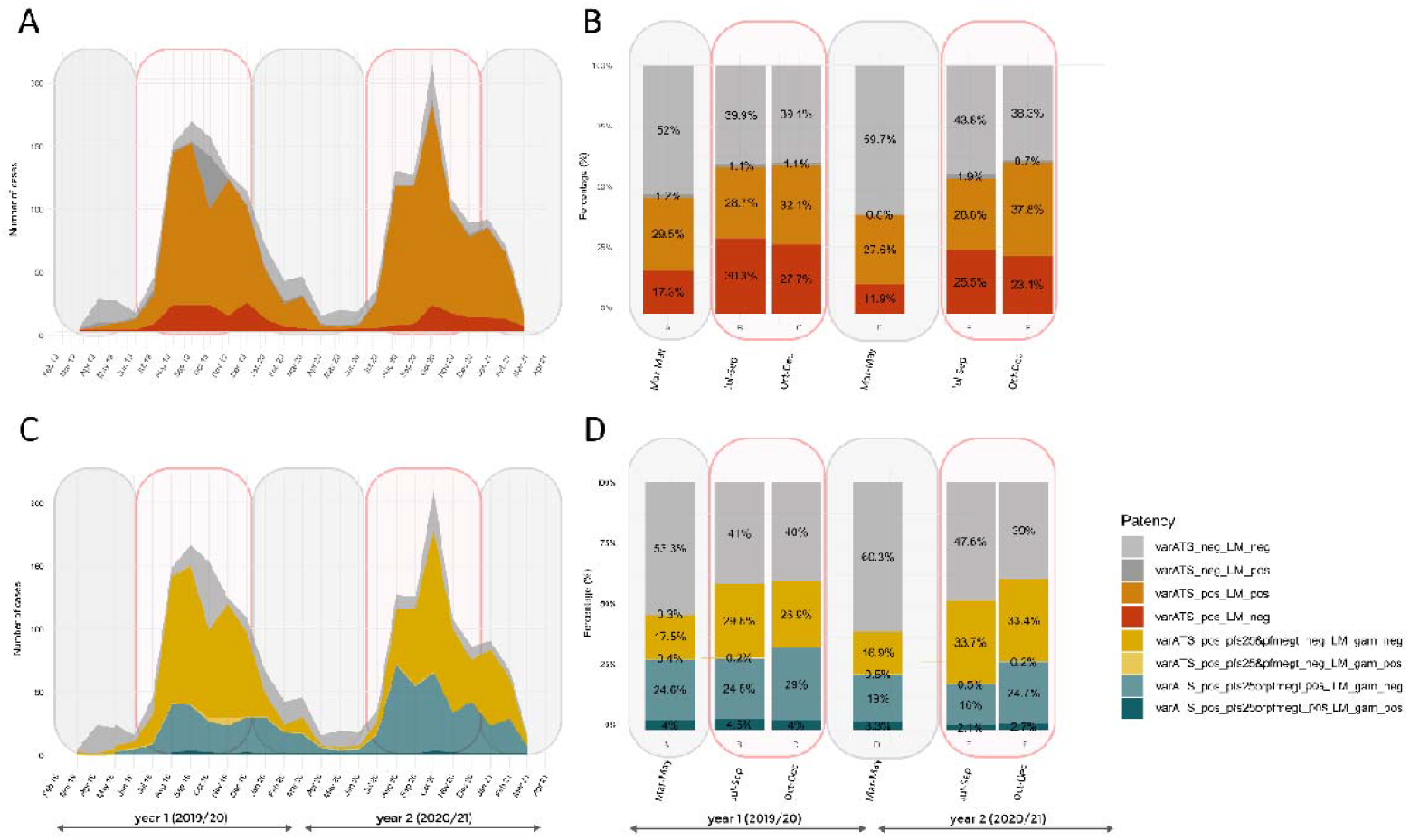
*P. falciparum* infection and gametocyte carriage dynamics during the study period. A) Number of suspected and confirmed malaria cases during the study period as detected by PCD. B) Percentage of asymptomatic infections per CSS during the ACD. B) Number of clinical cases carrying gametocytes during the study period as detected by PCD. D) Gametocyte carriage among asymptomatic cases per CSS; Time (months) is shown on the x-axis, with HTS months highlighted in red-lined boxes and LTS months in grey. The y-axis represents the number of cases in panel (A/C) and proportions in panel (B/D). Areas in (A) and bars in (B) illustrate the proportion of microscopic and sub-microscopic infections per month in (A) and CSS in (B). Areas (C) and bars (D), show the proportion of asexual infections (yellow), gametocyte carriers (green), sub-microscopic gametocyte carriers (light green), gametocytes detected by LM (dark green) per month in (C) and CSS in (D). ACD, active case detection; PCD, passive case detection, HTS, high transmission season, LTS, low transmission season, LM, light microscopy, neg, negative, pos, positive, CSS, cross-sectional survey.

The crude prevalence of asymptomatic infections ranged from 262/663 (39·5%) to 379/811 (46·7%) during LTS and from 324/596 (54·4%) to 345/544 (61·0%) during HTS (Figure 3B, yellow and red). During ACD sub-microscopic infections were more frequent during the HTS (695/2805 [24·8%]) than during LTS (235/1573 [14·9%]; χ^2^=57·713, df=1, *P*<0·0001).

In clinical cases (PCD), the number of gametocyte carriers followed the dynamic of clinical infections, peaking during HTS (Figure 3C, green). The percentage of asymptomatic infections carrying gametocytes was highest during LTS (223/367 [60·8%] in 2019; 147/261 [55·1%] in 2020), decreased at the beginning of HTS (190/384 [49·5%] in 2019; 103/298 [34·7%] in 2020) and increased again at the end of HTS (174/314 [55·1%] in 2019; 151/336 [44·9%] in 2020, χ^2^=56·206, df=5, *P*<0·0001). The percentage of the total population during ACD carrying gametocytes remained stable through the seasons (HTS 618/2296 [26·9%], LTS 370/1443 [25·6%], χ^2^=0·677, df=1, *P*=0·4106) (Figure 3D, green). The majority of gametocyte carriers were not detectable by LM (Figure 3D, dark green).

Among sub-microscopic cases, gametocyte carriage remained high during all CSS (40/164 [24·4%]-67/169 [39·6%]), although significantly lower than in microscopic cases (91/208 [43·8%]-185/245 [75·5%], χ^2^=191·69, df=1, *P*<0·0001) (Supplementary Figure 3).

The adjusted prevalence of asymptomatic infection was lower among participants with HbAS (Figure 4A), while gametocyte carriage among asymptomatic infections did not differ significantly by Hb genotypes (Figure 4B).

**Figure 4.**
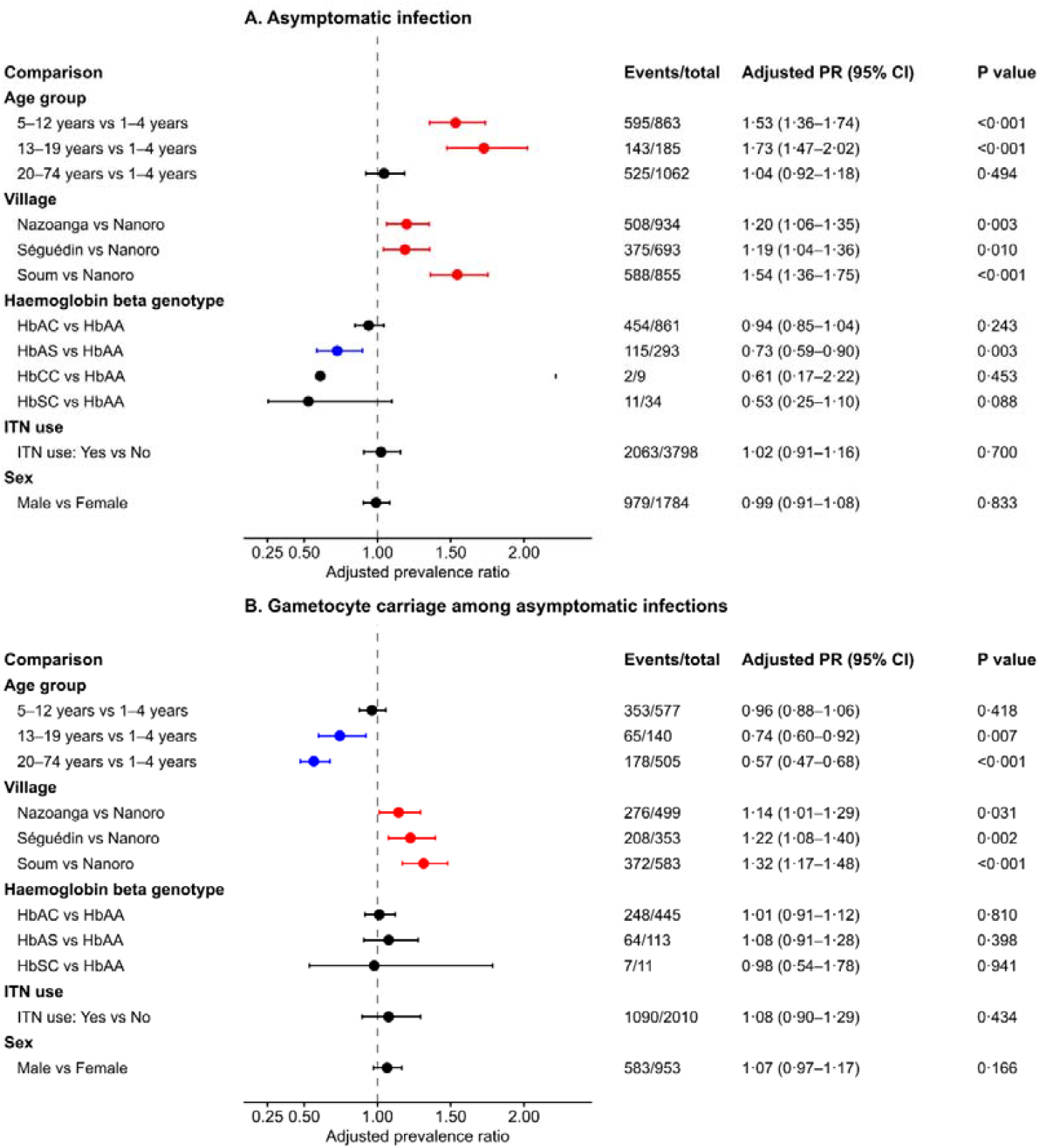
Adjusted prevalence ratios for factors associated with asymptomatic *P. falciparum* infection (A) and gametocyte carriage among asymptomatic infections (B). Estimates were derived from mixed-effects models adjusted for sex, age group, village, ITN use, and haemoglobin beta genotype, with random intercepts for participant ID and cross-sectional survey visit. Horizontal lines indicate 95% confidence intervals; the vertical dashed line indicates the null value, PR=1. Red indicates significantly higher adjusted prevalence, blue indicates significantly lower adjusted prevalence, and black indicates non-significant associations. HbCC was excluded from the gametocyte carriage model because this category was too sparse to estimate reliably.

Compared with children aged 1–4 years, the adjusted prevalence of asymptomatic *P. falciparum* infections was 53% higher in school-aged children and 73% higher in adolescents (Figure 4A and 5A). In contrast, the adjusted prevalence of gametocyte carriage among asymptomatic infections was 26% lower in adolescents and 43% lower in adults, while it was similar in school-aged children (Figure 4B).

Village of residence was also associated with both outcomes. Compared with Nanoro, the adjusted prevalence of asymptomatic infection was 20% higher in Nazoanga, 19% higher in Séguédin, and 54% higher in Soum (Figure 4A). For gametocyte carriage among asymptomatic infections, adjusted prevalence was also higher in all three villages compared with Nanoro (Figure 4B). These differences were consistent with the observed prevalence of microscopic and sub-microscopic asymptomatic infections across villages (Figure 5C).

**Figure 5.**
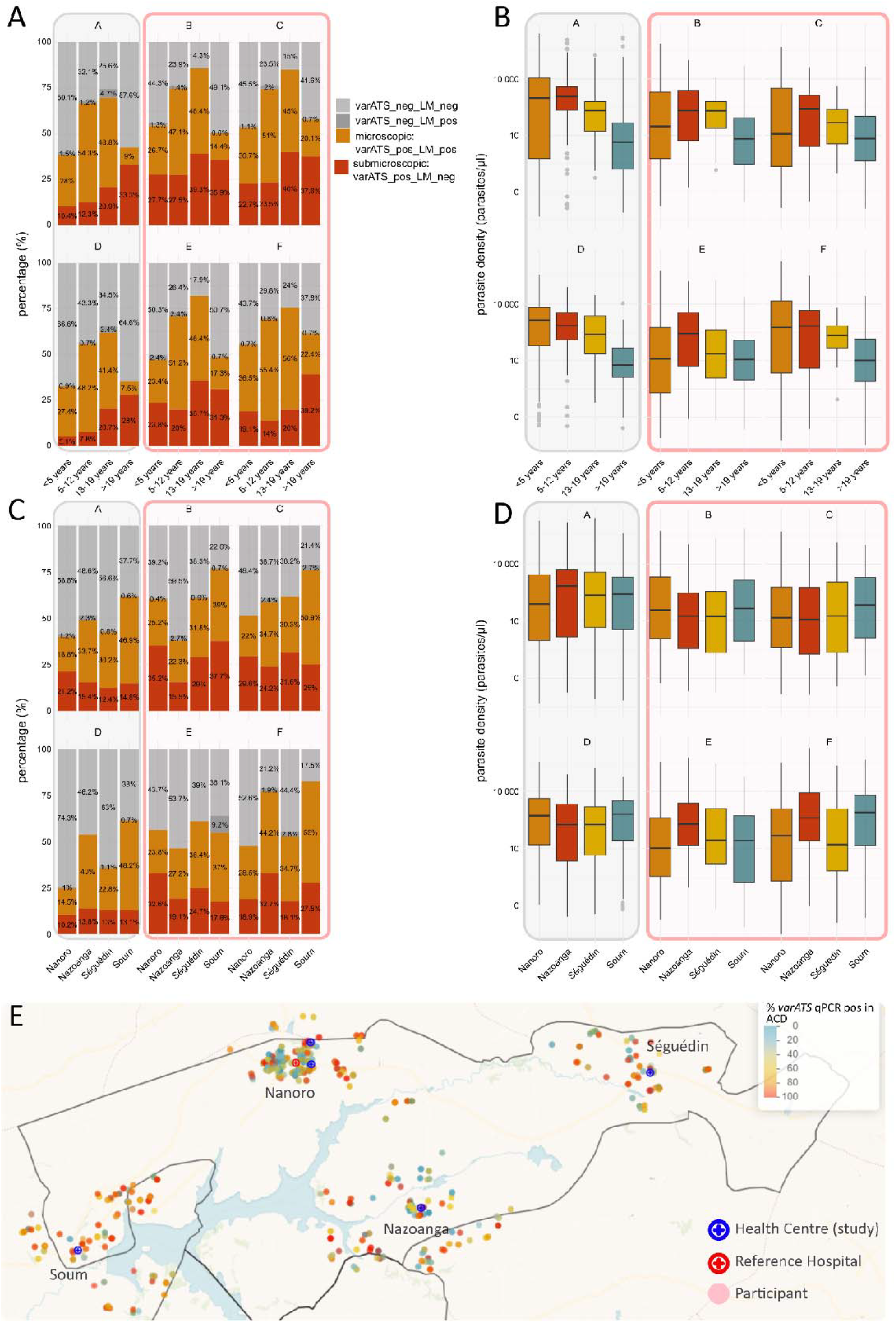
Variation of asymptomatic *P. falciparum* infections per village and age during the six cross-sectional surveys. Prevalence *P. falciparum* infections calculated as percentage of *varATS* qPCR and/or LM positive individuals per CSS per age group (panel A) or village (panel C). Parasites densities (panel B and D) per age group (panel B) or village (panel D) were obtained by interpolating cycle thresholds (Ct) using a standard curve prepared with titrated samples containing known numbers of infected erythrocytes diluted in whole blood (100,000–0.01 parasites/μL). Bars in (panel A & C) illustrate the proportion of microscopic and sub-microscopic infections per CSS per age group (A) or village (C). CSS conducted during the LTS in grey-lined boxes, HTS highlighted in red-lined boxes. Panel E) Individual frequency of asymptomatic carriage (*varATS* qPCR positives in ACD) across the study villages. Map showing the location of the study participant’s HH within the study area and the percentage of asymptomatic infections detected per individual during the 6 cross-sectional surveys (blue cross: study health centre, red cross: reference hospital, each dot represents participants’ HH). HTS, high transmission season, LTS, low transmission season, LM, light microscopy, neg, negative, pos, positive, CSS, cross-sectional survey. Basemap: CARTO Voyager (© CARTO), based on OpenStreetMap data (© OpenStreetMap contributors).

We mapped individual frequency of asymptomatic carriage (*varATS* qPCR positives in ACD) across the study villages (Figure 5E); correlations between individual frequency of asymptomatic carriage and distance to water body and study health centre were weak (health centre, R=0·15, *P*<0·001, water body, R=-0·09, *P*=0·02), with no correlation with gametocyte carriage (health centre, R=0·04, *P*=0·45, water body, R=0·02, *P*=0·729).

To further investigate the potential transmission contribution of individuals by age groups and transmission season, we applied thresholds to our of gametocytes/µL from a previous MFA study in Burkina Faso as a proxy to define an individual’s potential to infect mosquitoes (Supplementary Table 3).^17^ Gametocyte densities showed a bimodal pattern with a higher peak at low gam/µL values (around 1 gam/µl) and a lower peak at higher gam/µL values (around 100 gam/µl) (Figure 6A). Overall, the prevalence of potentially infectious individuals did not differ between asymptomatic and clinical infections (478/3668 [13.0%] and 286/2189 [13.0%] respectively, χ^2^ <0·0001, df=1, *P*=1). Transmission potential decreased markedly with age, being highest in children younger than five years (551/3321 [16·6%]) and school-aged children (182/1139 [16·0%]), and drastically lower in older individuals (16/208 [7·6%] and 15/1195 [1·3%] in adolescents and adults respectively) (χ^2^=197·33, df=3, *P*<0·0001).

**Figure 6.**
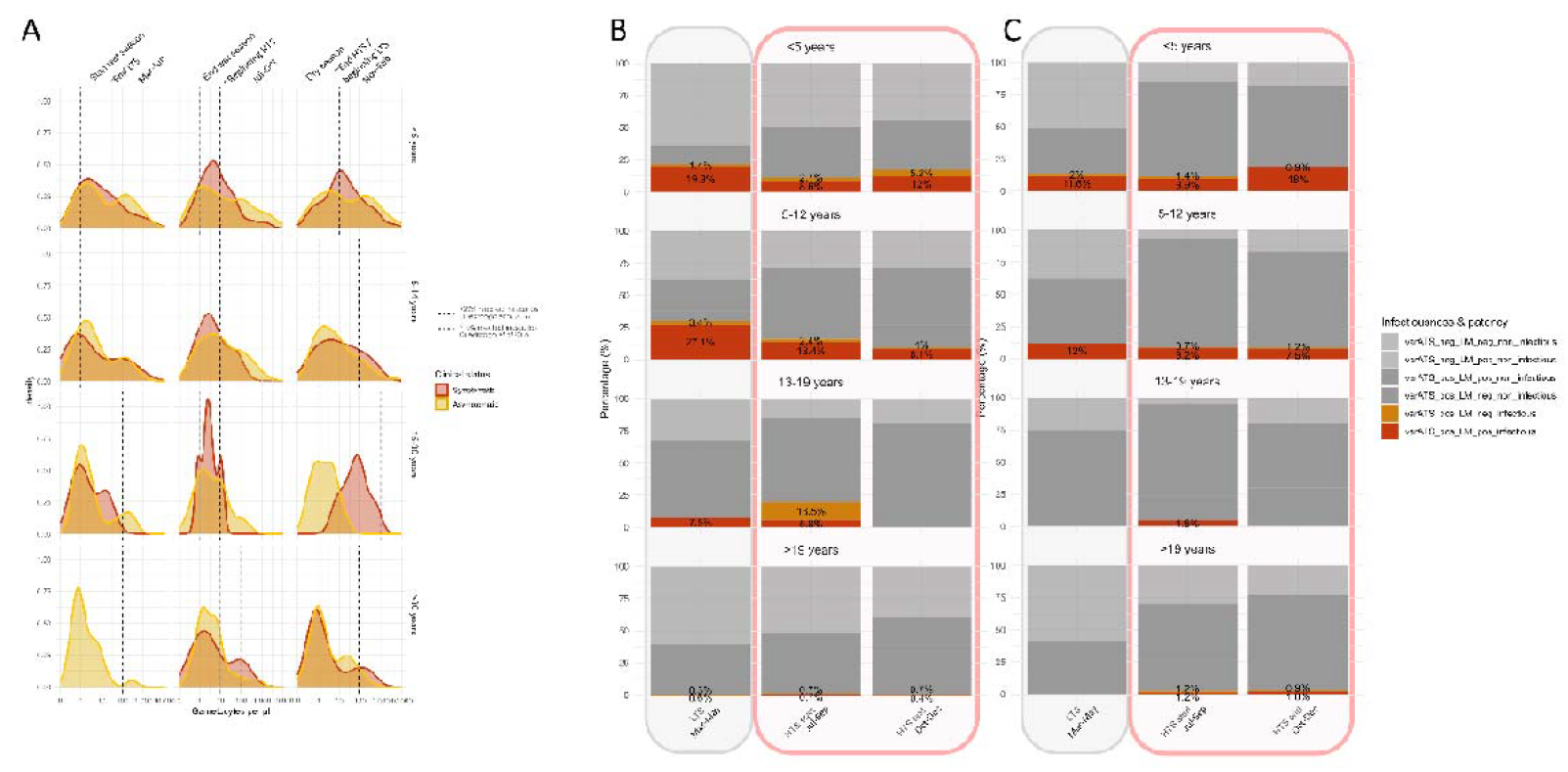
Infectious reservoir by age, season, clinical status. A) Density plot of gametocytes/µL µL based on *pfs25* and *pfmget* mRNA copy numbers per age group and season in asymptomatic and symptomatic individuals. Applied thresholds of infected mosquitoes are based on relationship between gametocytes/µL values and successful membrane feeding assay (MFA) results generated from Ouédraogo *et al*, 2016 (10). Thresholds of gametocytes/µL (gam/µL) to successfully infect over >10% (grey dashed line) or >20% of mosquitos (black dashed line) are differed by season and age group in their data (see also Supplementary Table 5). A study carried out in a similar study area (50-100 km from the study area of the current study). For comparability, age groups and seasons were aligned to age groups and seasons in Ouédraogo *et al* 2016: March-June = Start wet season, July-October = End wet season, November – February = Dry season. Only observations positive for gametocyte carriage are plotted. B/C Infectious reservoir as defined by the threshold of > 20% infected mosquitos by age group and season divided by microscopic (red) and sub-microscopic infections (orange) in asymptomatic (B) and symptomatic (C) individuals. HTS highlighted in red-lined boxes and LTS in grey. HTS, high transmission season, LTS, low transmission season, LM, light microscopy, neg, negative, pos, positive, CSS, cross-sectional survey.

Transmission potential peaked during the LTS in younger age groups (159/758 [20·8%] children younger than five years and 92/311 [29·6%] school-aged children) and declines at the end of the HTS (111/637 [17·4%] children younger than five years and 22/240 [9·2%] school-aged) (Figure 6B/C). Prevalences of infectious individuals were highest in Soum (262/1188 [22·1%]) and Nazoanga (173/1370 [12·6%]) compared to Nanoro (202/2348 [8·6%], χ^2^ = 126·21, df = 3, p-value < 0·001).

Longitudinal analysis over the study period of asymptomatic infections and clinical episodes showed that individuals in all age groups can remain asymptomatic without clinical episodes between consecutive seasons (Figure 7A in yellow). The highest asymptomatic persistence was observed in school-aged children and adolescents (in school-aged children in 2019 67/175 [38·3%] and 2020 50/169 [29.6%] of events emerging from LTS remain asymptomatic and 16/44 [36.4%] and 11/28 [39.3%] in adolescents, respectively). It is noteworthy that most clinical cases are reinfections of the same individual rather than emerging from asymptomatic infections (Figure 7A, in red, reinfections as seen as circular patterns). Of 706 total clinical cases in 2019, 185 had a clinical episode during the same HTS before [26·2%] and 131/709 [18·5%] in 2020. In 2019 only 58 clinical cases were asymptomatic infections in the LTS before [8·2%] and 32 [4·5%] in 2020 (Supplementary Table 4).

**Figure 7.**
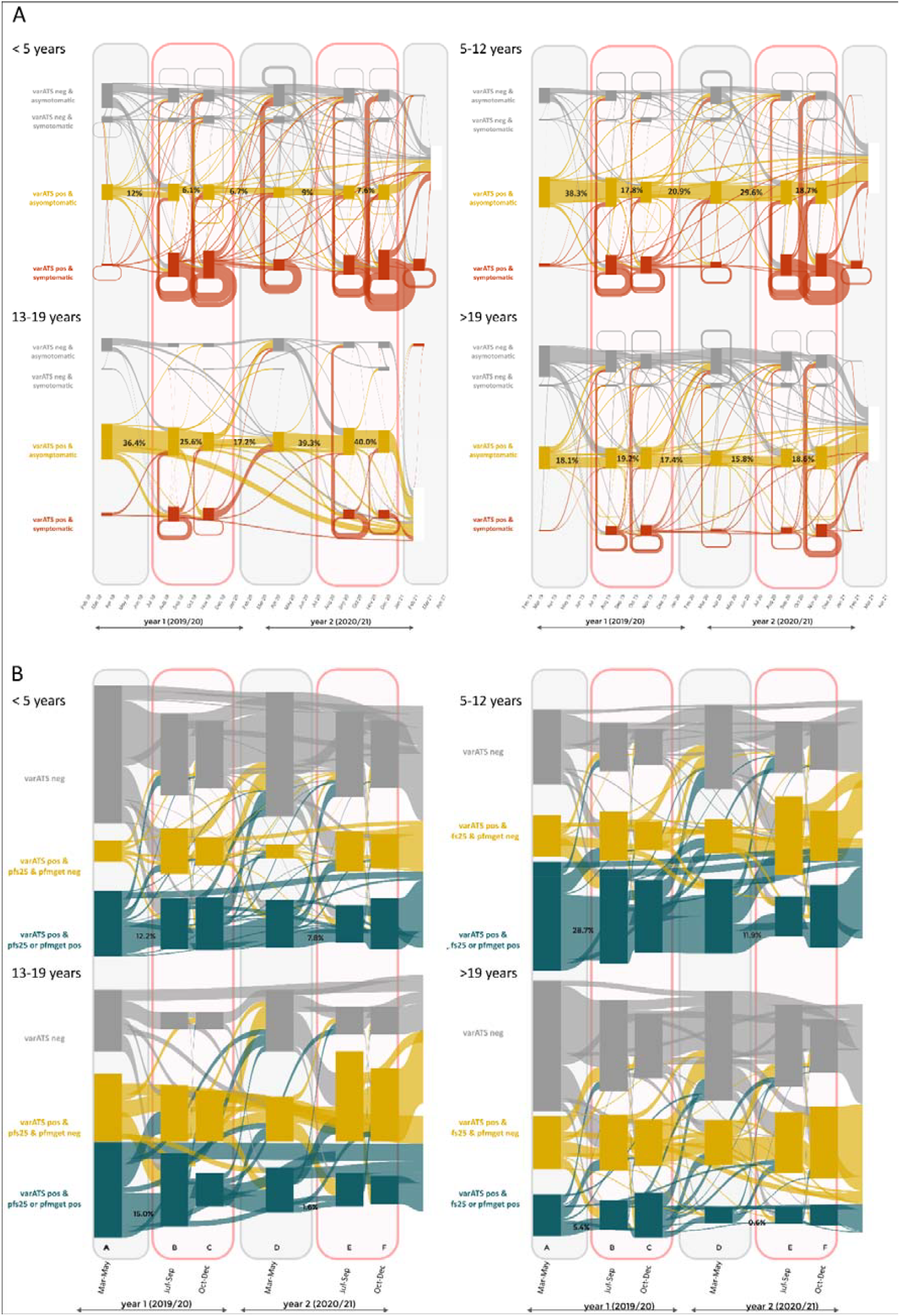
Dynamics of asymptomatic infections and symptomatic infections (A) and gametocyte carriage in ACD (B) cases per age groups and transmission seasons over the study period. Each panel represents an age group. X-axis is time (months); months in red lined boxes are HTS and grey are LTS. A) ACD and PCD data were pooled and categorized in seven periods, based on the 6 cross-sectional collections: LTS (March – June) 2019, 2020 and 2021, HTS start (July-August) and HTS end (September-December) 2019 and 2020. Y-axis shows proportions of *P. falciparum* negative infections (grey), asymptomatic asexual infections (yellow) and symptomatic asexual infections (red). B) Y axis shows proportions of *P. falciparum* negative (grey), asexual infections (yellow) and gametocyte carriers (green). Connecting lines depict portions of individuals shifting from one state to another or remaining in the same state between two periods (panel A) or CSS (panel B). HTS, high transmission season, LTS, low transmission season. ACD, active case detection, LM, light microscopy, neg, negative, pos, positive.

In the younger age group (<13 years old), a large percentage of participants remained gametocyte positive from one CSS to the next (Figure 7B, in green). Between LTS and HTS, the percentage of individuals who remained gametocyte positive was highest in school-aged children (45/157 [28·7%] in 2019 and 16/135 [11·9%] in 2020), followed by children younger than five years (47/384 [12·2%] in 2019 and 25/319 [7·8%] in 2020), and lowest in individuals in older age groups of adolescents (6/40 [15·0%] in 2019 and 1/27 [3·7%] in 2020) and adults (11/205 [5·4%] in 2019 and 1/156 [0·6%] in 2020).

## DISCUSSION

This study describes the dynamics of *P. falciparum* infections and gametocyte carriage in an all-age population across six cross-sectional surveys within a longitudinal cohort over two consecutive years in a highly seasonal transmission setting. Using molecular diagnostics, we identified a high prevalence of asymptomatic infections in the study population during both the LTS (39·5–46·7%) and the HTS (54·4–61·0%). Among these infections, the prevalence of gametocyte carriage-representing the infectious reservoir - was high during both LTS (55·1-60·8%) and HTS (34·7-55·1). However, less than half of these individuals (545/1232 [44·2%]) carried gametocytes at densities sufficient to infect mosquitoes (as estimated using an infectiousness threshold derived from the association between gametocyte density and mosquito infection success in membrane feeding assays conducted in Burkina Faso). To inform targeted transmission-reducing interventions, we further characterized the demographic and temporal features of this reservoir.

Age emerged as the strongest predictor of asymptomatic infection and gametocyte carriage, while school-aged children and adolescents were at highest risk for asymptomatic infection, children younger than 13 years were at highest risk for gametocyte carriage and correspondingly had the highest estimated transmission potential. These findings are consistent with previous studies across sub-Saharan Africa, including Burkina Faso.^4,6,7^

While we see a similar transmission potential in clinical and asymptomatic individuals, clinical cases are more likely to be detected and treated under current “test-and-treat” strategies. We investigated whether asymptomatic infections likely develop into clinical episodes and thus get treated. Only a small percentage (4·5-8·2%) of asymptomatic infections detected during LTS progressed to clinical disease in the subsequent HTS. This contrasts with studies reporting increased risk of clinical episodes following asymptomatic infection,^18,19^ but supports evidence suggesting partial protection against symptomatic disease.^20–22^

Long-lasting asymptomatic infections (those persisting from one season to the next) were especially common in school-aged children and adolescents (12·4 and 20·9 % of persistent asymptomatic carriage from one season to the next in these age groups, respectively, compared to 8·8 and 3·7 % in adults and children younger than five, respectively). School-aged children also accounted for the largest percentage of persistent gametocyte carriers (12·4 % persistent asymptomatic gametocyte carriage from one CSS to the next, compared to 7·6, 5·1, and 2·4 % in adolescents, children younger than five and adults respectively). With the observed transmission potential peaking during the LTS, our data highlight the transition from LTS to HTS —when mosquito populations re-emerge and reinfection is required to reinitiate transmission— as a critical window for transmission-reducing interventions. We observed marked spatial heterogeneity, with the highest infection risk and transmission potential in Soum, a site located near a large dam. These findings emphasise the importance of micro-epidemiological variation and the need for context specific data to guide intervention strategies. The correlations we observe between asymptomatic infection and distance to health centre point towards the need to increase efforts to reach remote populations. Understanding the drivers and consequences of spatial heterogeneity will help identify transmission hotspots and improve targeted control strategies.^23^

Most infectious individuals were detectable by LM, suggesting that standard diagnostic tools can capture a substantial fraction of the infectious reservoir. Although in our longitudinal analyses reinfection cannot be excluded in the absence of genotyping data, repeated detection of asymptomatic infection and gametocyte carriage in the same individuals indicates sustained individual-level contribution to transmission. Although gametocyte density is strongly associated with mosquito infection rates,^7^ infectiousness is also influenced by host-related factors, including mosquito exposure, attractiveness, and immunity.^5,24^ While membrane feeding assays were not conducted in this study, applying a previously established density–infectivity relationship provided a context-specific proxy for transmission potential. We identified a bimodal distribution of gametocyte densities, with a major peak at low densities (~1 gametocyte/μL) and a second peak at higher densities (~100 gametocytes/μL). Given that transmission is more likely at densities exceeding 1 gametocyte/μL—and particularly above 10 gametocytes/μL^25^—the higher-density peak, most prominent in LTS season, likely represents the core infectious reservoir.

Taken together, these findings support targeted interventions focusing on school-aged children, who represent the largest and most persistent infectious reservoir and yet are not routinely targeted by current strategies. Interventions deployed at the end of the LTS may be particularly effective. Transmission-blocking drug regimens combining artemisinin combination therapies with single dose primaquine, could be particularly impactful in limiting the resurgence of transmission.^26,27^ Since we observe that only half of ITN used by the study population were in good condition and that the percentage of participants who slept under an ITN was lowest among school-aged children and adolescents, targeted ITN distribution and sensitisation on their use in those groups might proof impactful.

Preventive strategies, including intermittent preventive treatment in school-aged children (IPTsc)^28,29^ or SMC to school-aged children^30^ may decrease the long lasting asymptomatic reservoir. The potential impact of malaria vaccines on transmission dynamics remains to be established.

## Supporting information

STROBE statement

## Contributors

DFO (supervision - field work, investigation, conducting experiments -- sample pre-processing, writing - review & editing), VK (data curation, formal analysis, visualization, writing - original draft, review and editing), TR (data curation, formal analysis, writing - review & editing), PG (validation, methodology, conducting experiments - lab procedures including qPCR-*pfs25, pfmget*, gene expression, writing - review & editing), AM (investigation - clinician, writing - review & editing), AMM (methodology, validation, conducting experiments, writing - review & editing), YDB (methodology, conducting experiments, writing - review & editing), ACP (methodology, conducting experiments, writing - review & editing), NRR (methodology, conducting experiments, writing - review & editing), ES (methodology, conducting experiments, writing – review & editing), AA (formal analysis, writing - review & editing), RO (conducting experiments- sample processing, qPCR, writing - review & editing), KD (participants’ recruitment, supervision, study implementation, writing - review & editing), MAS (supervision - lead clinician, coordinating clinical aspects, writing - review & editing), EH (supervision, writing - review & editing), AS (supervision, writing - review & editing), HS (supervision, writing - review & editing), HT (supervision, writing - review & editing), ARU (funding acquisition, supervision, methodology, conceptualization - study design, investigation, project administration, writing - original draft, review and editing), HMN (funding acquisition, supervision, methodology, conceptualization - study design, investigation - coordination field work, project administration, writing - original draft, review and editing)

## Data sharing

Due to the sensitive nature of the data, the authors are unable to share the data directly. Requests to access the data can be made to ITM’s contact point for data access. All requests will be reviewed for approval by ITMs Data Access Committee. After approval, data sharing will be managed by the same committee.

## Acknowledgements

We are grateful to the Nanoro health district community for their participation in this study, and all the staff of the Clinical Research Unit of Nanoro.

This study was funded by the Belgium Directorate General for Development Cooperation (DGD) through the collaborative framework agreement 4 and 5 (FA4-FA5–DGD programme) between Clinical Research Unit of Nanoro (CRUN, Burkina Faso) and the Institute of Tropical Medicine in Antwerp, Belgium.

Research Foundation Flanders supported ES and YDB through FWO PhD-SB Fellowships (ES: 1SC5522N; YDB: 1S74321N) and VK through a “Research project grant” (G067823N).

## References

1 Portugaliza HP, Natama HM, Guetens P, et al. Plasmodium falciparum sexual conversion rates can be affected by artemisinin-based treatment in naturally infected malaria patients. EBioMedicine 2022; 83: 104198.

2 Andolina C, Ramjith J, Rek J, et al. Plasmodium falciparum gametocyte carriage in longitudinally monitored incident infections is associated with duration of infection and human host factors. Sci Rep 2023; 13: 7072.

3 World malaria report 2025. https://www.who.int/teams/global-malaria-programme/reports/world-malaria-report-2025 (accessed May 6, 2026).

4 Rek J, Blanken SL, Okoth J, et al. Asymptomatic School-Aged Children Are Important Drivers of Malaria Transmission in a High Endemicity Setting in Uganda. J Infect Dis 2022; 226: 708–13.

5 Stone W, Gonçalves BP, Bousema T, Drakeley C. Assessing the infectious reservoir of falciparum malaria: past and future. Trends Parasitol 2015; 31: 287–96.

6 Ouattara SM, Ouattara D, Badoum ES, et al. Malariometric Indices in the Context of Seasonal Malaria Chemoprevention in Children Aged 1.5 to 12 Years during the Period of High Malaria Transmission in the Suburban Area of Banfora, Burkina Faso. Trop Med Infect Dis 2023; 8: 442.

7 Andolina C, Rek JC, Briggs J, et al. Sources of persistent malaria transmission in a setting with effective malaria control in eastern Uganda: a longitudinal, observational cohort study. Lancet Infect Dis 2021; 21: 1568–78.

8 Barry A, Bradley J, Stone W, et al. Higher gametocyte production and mosquito infectivity in chronic compared to incident Plasmodium falciparum infections. Nat Commun 2021; 12: 2443.

9 Derra K, Rouamba E, Kazienga A, et al. Profile: Nanoro Health and Demographic Surveillance System. Int J Epidemiol 2012; 41: 1293–301.

10 Drakeley C, Abdulla S, Agnandji ST, et al. Longitudinal estimation of Plasmodium falciparum prevalence in relation to malaria prevention measures in six sub-Saharan African countries. Malar J 2017; 16: 433.

11 Natama HM, Ouedraogo DF, Sorgho H, et al. Diagnosing congenital malaria in a high-transmission setting: clinical relevance and usefulness of P. falciparum HRP2-based testing. Sci Rep 2017; 7: 2080.

12 Casas-Vila N, Pickford AK, Portugaliza HP, Tintó-Font E, Cortés A. Transcriptional Analysis of Tightly Synchronized Plasmodium falciparum Intraerythrocytic Stages by RT-qPCR. Methods Mol Biol Clifton NJ 2021; 2369: 165–85.

13 Wampfler R, Mwingira F, Javati S, et al. Strategies for Detection of Plasmodium species Gametocytes. PLOS ONE 2013; 8: e76316.

14 Wang CYT, Ballard E, Llewellyn S, et al. Assays for quantification of male and female gametocytes in human blood by qRT-PCR in the absence of pure sex-specific gametocyte standards. Malar J 2020; 19: 218.

15 Wittwer CT, Reed GH, Gundry CN, Vandersteen JG, Pryor RJ. High-resolution genotyping by amplicon melting analysis using LCGreen. Clin Chem 2003; 49: 853–60.

16 Bousema T, Drakeley C. Epidemiology and infectivity of Plasmodium falciparum and Plasmodium vivax gametocytes in relation to malaria control and elimination. Clin Microbiol Rev 2011; 24: 377–410.

17 Ouédraogo A, Gonçalves BP, Gnémé A, et al. Dynamics of the Human Infectious Reservoir for Malaria Determined by Mosquito Feeding Assays and Ultrasensitive Malaria Diagnosis in Burkina Faso. J Infect Dis 2016; 213: 90–9.

18 Fogang B, Lellouche L, Ceesay S, et al. Asymptomatic Plasmodium falciparum carriage at the end of the dry season is associated with subsequent infection and clinical malaria in Eastern Gambia. Malar J 2024; 23: 22.

19 Sumner KM, Mangeni JN, Obala AA, et al. Impact of asymptomatic Plasmodium falciparum infection on the risk of subsequent symptomatic malaria in a longitudinal cohort in Kenya. eLife 2021; 10: e68812.

20 Males S, Gaye O, Garcia A. Long-term asymptomatic carriage of Plasmodium falciparum protects from malaria attacks: a prospective study among Senegalese children. Clin Infect Dis Off Publ Infect Dis Soc Am 2008; 46: 516–22.

21 Portugal S, Tran TM, Ongoiba A, et al. Treatment of Chronic Asymptomatic Plasmodium falciparum Infection Does Not Increase the Risk of Clinical Malaria Upon Reinfection. Clin Infect Dis 2017; 64: 645–53.

22 Sondén K, Doumbo S, Hammar U, et al. Asymptomatic Multiclonal Plasmodium falciparum Infections Carried Through the Dry Season Predict Protection Against Subsequent Clinical Malaria. J Infect Dis 2015; 212: 608–16.

23 Stresman GH, Giorgi E, Baidjoe A, et al. Impact of metric and sample size on determining malaria hotspot boundaries. Sci Rep 2017; 7: 45849.

24 Dantzler KW, Ma S, Ngotho P, et al. Naturally acquired immunity against immature Plasmodium falciparum gametocytes. Sci Transl Med 2019; 11: eaav3963.

25 Gonçalves BP, Kapulu MC, Sawa P, et al. Examining the human infectious reservoir for Plasmodium falciparum malaria in areas of differing transmission intensity. Nat Commun 2017; 8: 1133.

26 John CC. Primaquine plus artemisinin combination therapy for reduction of malaria transmission: promise and risk. BMC Med 2016; 14: 65.

27 Mahamar A, Vanheer LN, Smit MJ, et al. Artemether–lumefantrine–amodiaquine or artesunate– amodiaquine combined with single low-dose primaquine to reduce Plasmodium falciparum malaria transmission in Ouélessébougou, Mali: a five-arm, phase 2, single-blind, randomised controlled trial. Lancet Microbe 2025; 6. DOI:10.1016/j.lanmic.2024.100966.

28 Cohee LM, Opondo C, Clarke SE, et al. Preventive malaria treatment among school-aged children in sub-Saharan Africa: a systematic review and meta-analyses. Lancet Glob Health 2020; 8: e1499–511.

29 Makenga G, Baraka V, Francis F, et al. Effectiveness and safety of intermittent preventive treatment with dihydroartemisinin-piperaquine or artesunate-amodiaquine for reducing malaria and related morbidities in schoolchildren in Tanzania: a randomised controlled trial. Lancet Glob Health 2023; 11: e1277–89.

30 Yaro JB, Tiono AB, Ouedraogo A, et al. Risk of Plasmodium falciparum infection in south-west Burkina Faso: potential impact of expanding eligibility for seasonal malaria chemoprevention. Sci Rep 2022; 12: 1402.

