## Supplementary figures and tables for "Dynamics of *Plasmodium falciparum* asymptomatic asexual and sexual stages across transmission seasons in a rural and high-transmission setting in Burkina Faso: a two-year longitudinal cohort study with cross-sectional surveys"

**Supplementary Figures**

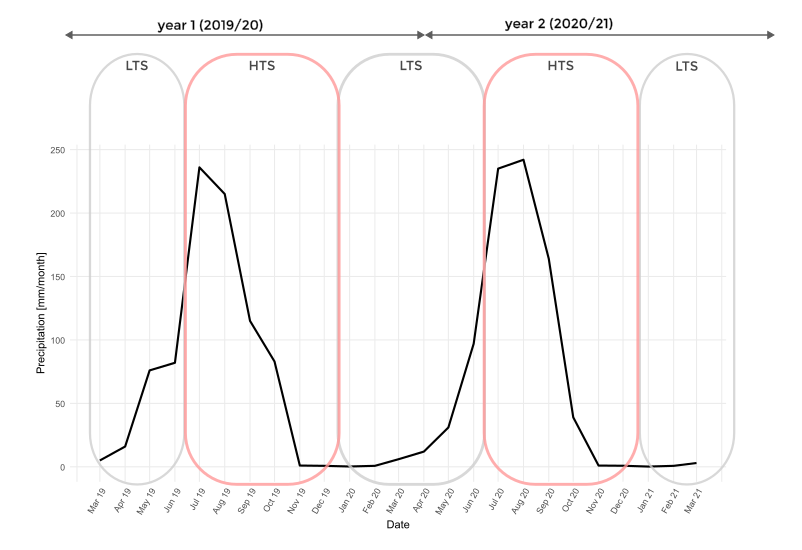

**Supplementary Figure 1. Precipitation in the study area during the study period.** X-axis is time (months); months in red lined boxes are high transmission season (HTS) and grey are low transmission season (LTS). Y-axis shows precipitation in mm/month based on “Climate Hazards Group InfraRed Precipitation with Station Data” (CHIRPS) [Funk 2015] . HTS, high transmission season, LTS, low transmission season.

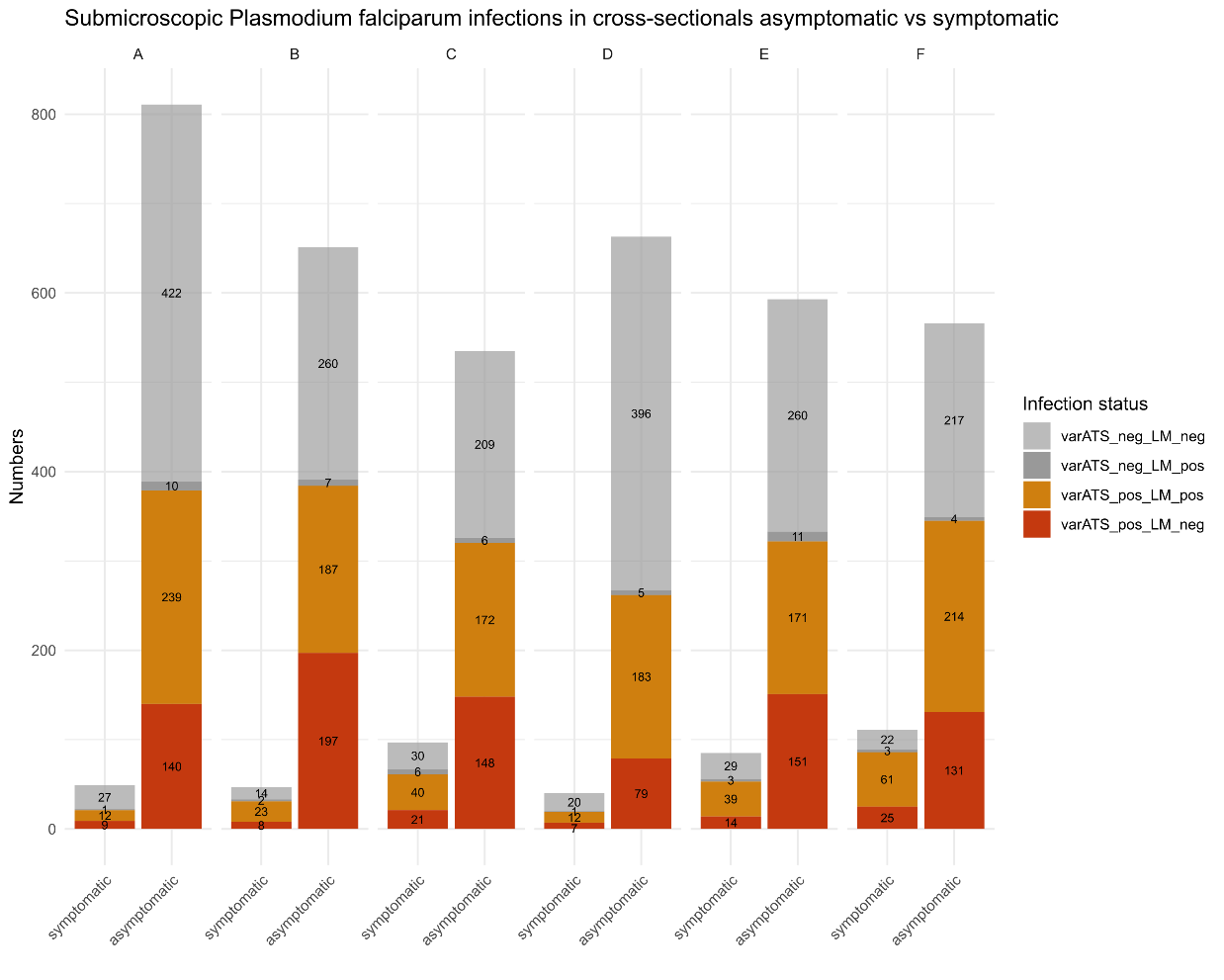

**Supplementary Figure 2. Number of asymptomatic and symptomatic microscopic and sub microscopic infections in ACD**. ACD, active case detection.

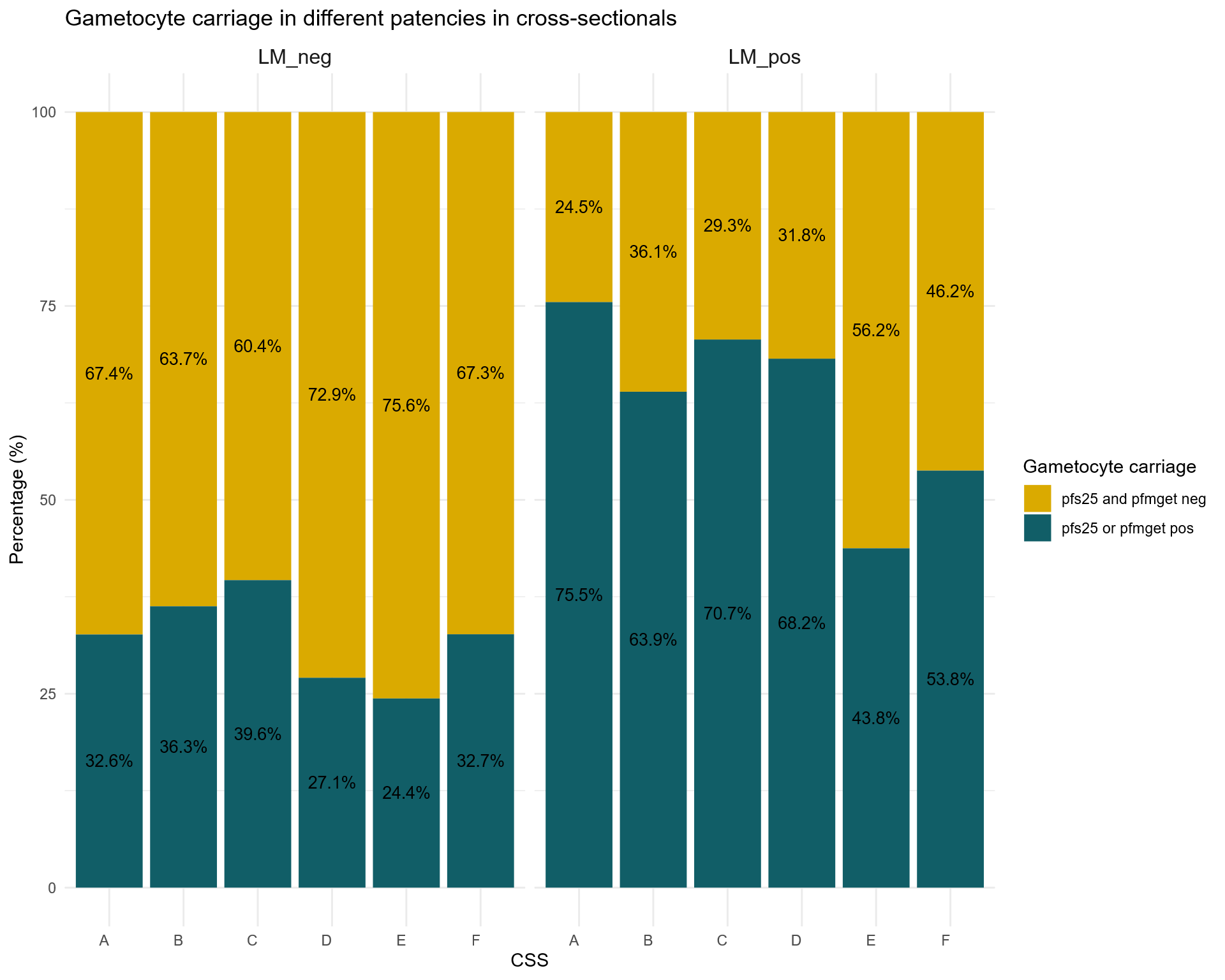

**Supplementary Figure 3. Asymptomatic gametocyte carriage among microscopic and sub microscopic cases.** LM, light microscopy, neg, negative, pos, positive, CSS, cross-sectional survey.

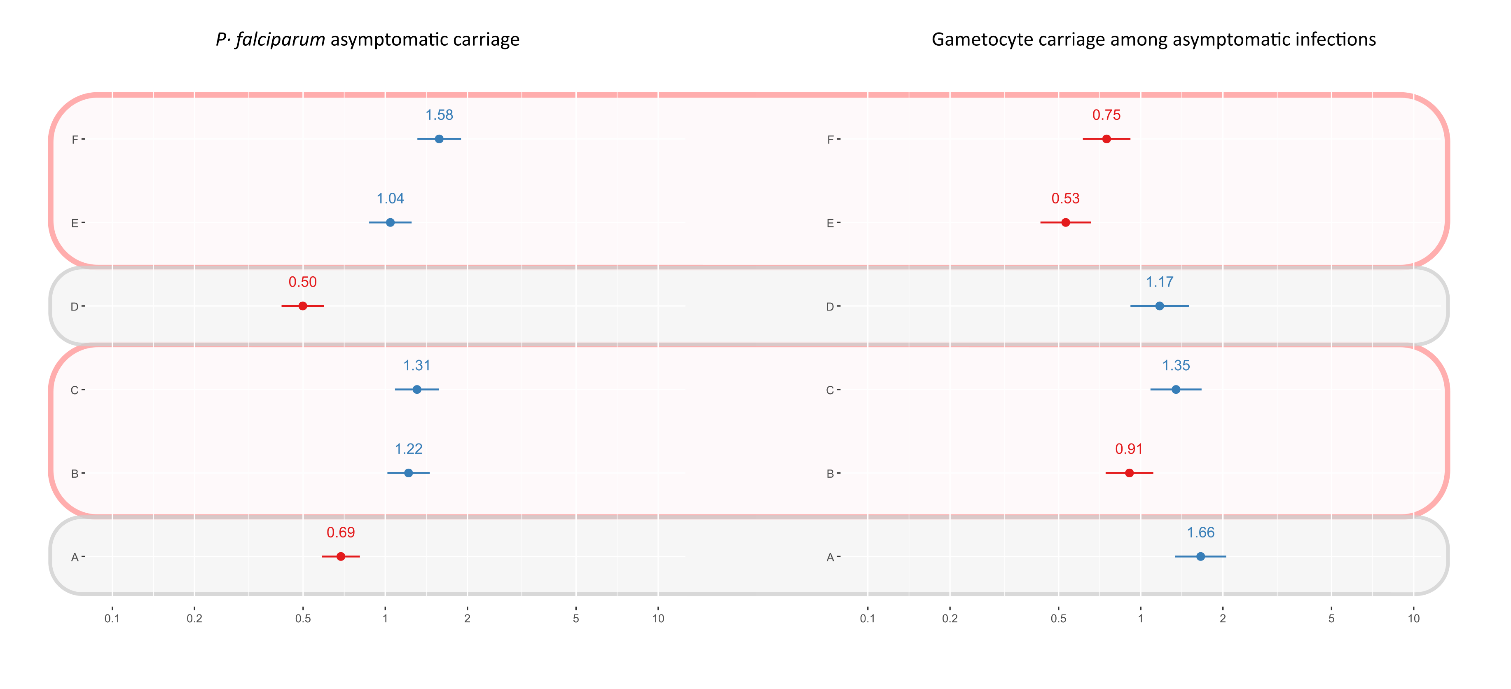

**Supplementary Figure 4 . Random effects of GLMM model for *P. falciparum* asymptomatic infection (panel A) and gametocyte carriage among asymptomatic infections (panel B) per CSS.** The odds of asymptomatic infection were lower during the CSS carried out during the LTS (grey boxes) compared to HTS (redlined boxes). The odds of gametocyte carriage in asymptomatic infections were higher during the CSS carried out during the LTS compared to HTS, apart from CSS C showing higher odds. X axis indicates the odds ratio (OR) and Y axis shows the CSS. HTS, high transmission season, LTS, low transmission season, CSS, cross-sectional survey.

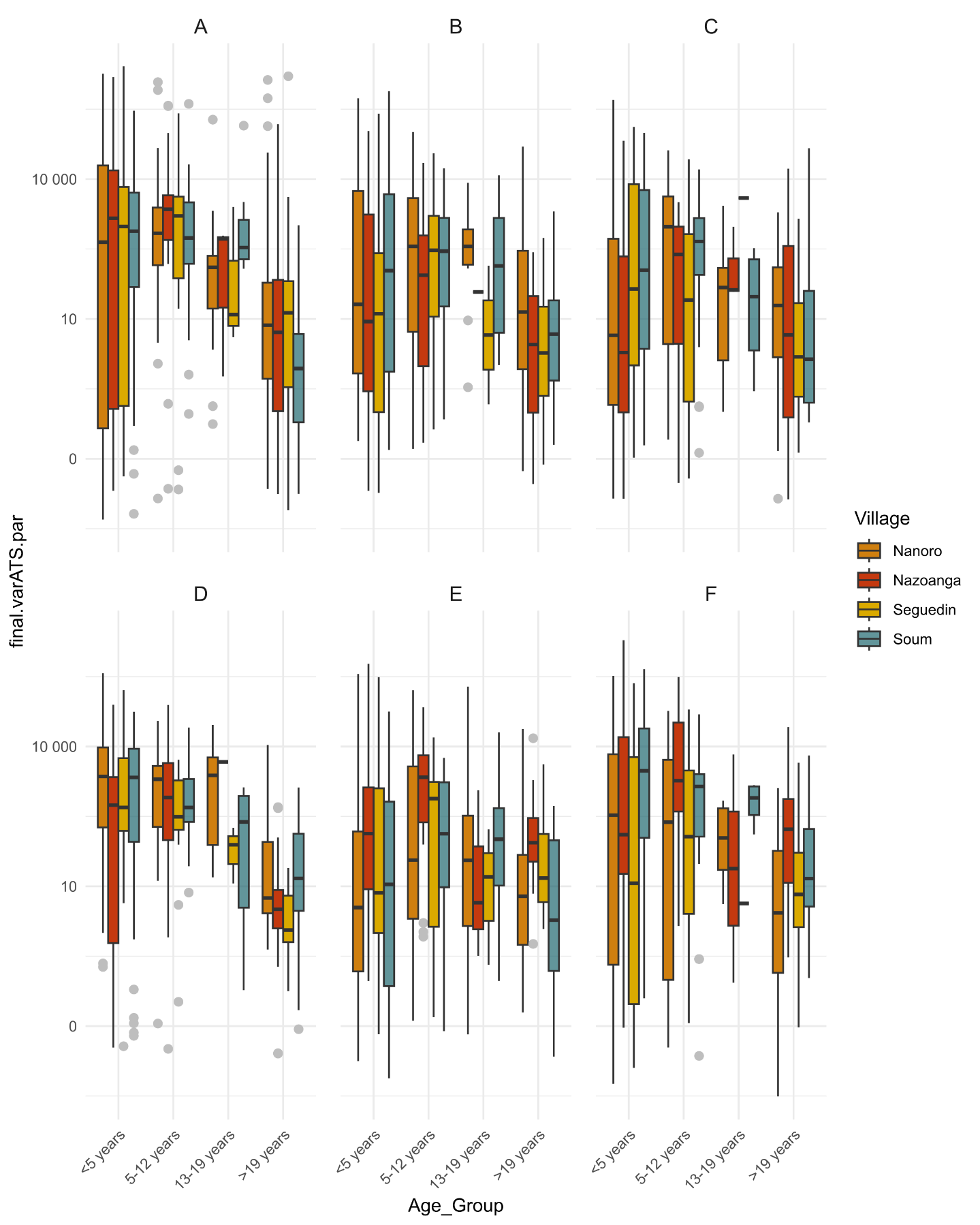

**Supplementary Figure 5. Parasite density in asymptomatic infection per village and age group.**

**Supplementary Tables**

| **Supplementary Table 1**. **Overview of primers used for the detection of *P. falciparum* asexual and sexual stages in this study.** | | | |
| --- | --- | --- | --- |
| **Name** | **Sequence 5’—3’** | **Target gene** | **Samples analyzed** |
| qPCR for the detection of *P. falciparum* asexual stage | | | |
| *varATS* Fw | 5’ – CCC ATA CAC AAC CAA YTG GA – 3’ | *varATS* | All |
| *varATS* Rev | 5’ – TTC GCA CAT ATC TCT ATG TCT ATC T – 3’ |  |  |
| qPCR for the detection of *P. falciparum* sexual stage | | | |
| *Pfmget* Fw | 5’ – GGTCCAAATATAAAATCC TGTTC – 3’ | *Pfmget* | *varATS* qPCR positive |
| *Pfmget* Rev | 5’ – TGTGTAACGTATGATTCATTTTC – 3’ |  |  |
| *Pfs25* Fw | 5’ – GAAATCCCGTTTCATACGCTTG – 3’ | *Pfs25* | *varATS* qPCR positive |
| *Pfs25* Rev | 5’ – AGTTTTAACAGGATTGCTTGTATCTAA – 3’ |  |  |

| **Supplementary Table 2. Software and databases used.** | | |
| --- | --- | --- |
| **Purpose** | **Name/URL** | **Reference** |
| Data curation | RedCap | [1, 2] |
| Data curation and analysis | R 4.4.1 (2024-06-14 ucrt) | [3] |
| Data curation analysis, visualization | geosphere, sp, raster, sf, tidyr, dplyr, lubridate, scales, readr, ggplot2, plotly, leaflet, gtsummary, binom, lme4, broom.mixed, performance, sjPlot, DHARMa, | [4-21] |
| Waterbody information | <https://diva-gis.org/data.html>. |  |
| Geospatial visualization | CARTO Voyager based on OpenStreetMap data | © CARTO, © OpenStreetMap contributors |
| Administrative boundary: | GADM | [22] |
| Visualization | Inkscape, version 1.2 | 2022 Inkscape Developers |

| **Supplementary Table 3· Summary of the percentage of individuals with gametocytes/µL levels that would be infectious to mosquitoes per age groups and transmission seasons** | | | | | | | |
| --- | --- | --- | --- | --- | --- | --- | --- |
|  |  |  |  | **percentage of individuals with gam/µL > threshold per group** | | | |
| **Age group** | **Season** | **Clinical status** | **n_total** | **1** | **10** | **100** | **1000** |
| <5 years | Start wet season | symptomatic | 176 | 17·05 | 9·09 | 2·84 | 1·14 |
|  |  | asymptomatic | 703 | 21·34 | 12·09 | 7·25 | 1·42 |
|  | End wet season | symptomatic | 774 | 30·88 | 12·92 | 3·10 | 1·16 |
|  |  | asymptomatic | 687 | 20·09 | 12·08 | 6·84 | 2·47 |
|  | Dry season | symptomatic | 593 | 31·70 | 20·57 | 7·76 | 1·52 |
|  |  | asymptomatic | 417 | 27·58 | 17·51 | 11·75 | 3·36 |
| 5-14 years | Start wet season | symptomatic | 36 | 16·67 | 11·11 | 5·56 | 0·00 |
|  |  | asymptomatic | 317 | 29·34 | 11·99 | 5·68 | 0·32 |
|  | End wet season | symptomatic | 230 | 25·65 | 9·57 | 1·74 | 0·43 |
|  |  | asymptomatic | 328 | 29·88 | 16·16 | 6·40 | 0·61 |
|  | Dry season | symptomatic | 131 | 25·19 | 14·50 | 5·34 | 0·00 |
|  |  | asymptomatic | 171 | 30·41 | 15·79 | 5·26 | 0·58 |
| 15-30 years | Start wet season | symptomatic | 12 | 16·67 | 8·33 | 0·00 | 0·00 |
|  |  | asymptomatic | 148 | 10·14 | 3·38 | 2·03 | 0·00 |
|  | End wet season | symptomatic | 59 | 11·86 | 3·39 | 0·00 | 0·00 |
|  |  | asymptomatic | 147 | 14·97 | 6·80 | 1·36 | 0·00 |
|  | Dry season | symptomatic | 28 | 21·43 | 17·86 | 7·14 | 0·00 |
|  |  | asymptomatic | 93 | 13·98 | 2·15 | 0·00 | 0·00 |
| >30 years | Start wet season | symptomatic | 21 | 0·00 | 0·00 | 0·00 | 0·00 |
|  |  | asymptomatic | 255 | 6·27 | 0·78 | 0·39 | 0·00 |
|  | End wet season | symptomatic | 106 | 10·38 | 3·77 | 1·89 | 0·00 |
|  |  | asymptomatic | 265 | 10·19 | 1·89 | 0·75 | 0·00 |
|  | Dry season | symptomatic | 64 | 7·81 | 3·13 | 1·56 | 0·00 |
|  |  | asymptomatic | 142 | 12·68 | 4·93 | 1·41 | 0·00 |
| Cells in grey >20% infected mosquitos cut off Ouédraogo 2016·Gam, gametocyte | | | | | | | |

| **Supplementary Table 4· Source of clinical malaria episodes during the high transmission season, based on Sankey diagram shown in Figure 7A·** | | | | | |
| --- | --- | --- | --- | --- | --- |
| **Year** | **Number of *varATS* pos asymptomatics during LTS becoming *varATS* pos symptomatic in HTS** | **%** | **Number of *varATS* pos symptomatic during HTS being *varATS* pos symptomatic during HTS before** | **%** | **all *varATS* pos symptomatics per HTS season** |
| **2019** | 58 | 8·2 | 185 | 26·2 | 706 |
| **2020** | 32 | 4·5 | 131 | 18·5 | 709 |
| HTS, high transmission season, LTS, low transmission season, pos, positive, | | | | | |

**Search strategy research in context**

**Web of Science**

TS=("Plasmodium falciparum" OR "P. falciparum" OR "falciparum malaria")

AND

TS=("asymptomatic malaria" OR "asymptomatic infection" OR "subclinical malaria" OR "clinical malaria" OR "symptomatic malaria" OR "uncomplicated malaria" OR "asymptomatic parasitaemia" OR "asymptomatic parasitemia" OR "asymptomatic carriage")

AND

TS=(gametocyte* OR gametocytaemia OR gametocytemia OR "sexual stage" OR "asexual stage" OR "asexual parasitaemia" OR "asexual parasitemia" OR "parasite density" OR "parasite carriage" OR "parasite prevalence")

AND

TS=("cross-sectional" OR "longitudinal" OR "prospective cohort" OR "cohort study" OR "follow-up study" OR "repeated survey*" OR "repeat cross-section*")

AND

TS=("2 year*" OR "two year*" OR "24 month*" OR "3 year*" OR "three year*" OR "4 year*" OR "four year*" OR "5 year*" OR "long-term follow*")

**PubMed**

("Plasmodium falciparum"[MeSH Terms] OR "Plasmodium falciparum"[tiab] OR "P. falciparum"[tiab] OR "falciparum malaria"[tiab])

AND

("asymptomatic malaria"[tiab] OR "asymptomatic infection"[tiab] OR "subclinical malaria"[tiab] OR "clinical malaria"[tiab] OR "symptomatic malaria"[tiab] OR "uncomplicated malaria"[tiab] OR "asymptomatic parasitaemia"[tiab] OR "asymptomatic parasitemia"[tiab])

AND

("Protozoan Life Cycle"[MeSH] OR gametocyte*[tiab] OR gametocytaemia[tiab] OR gametocytemia[tiab] OR "sexual stage"[tiab] OR "asexual stage"[tiab] OR "asexual parasitaemia"[tiab] OR "asexual parasitemia"[tiab] OR "parasite density"[tiab] OR "parasite prevalence"[tiab] OR "parasite carriage"[tiab])

AND

(("Cross-Sectional Studies"[MeSH] OR "cross-sectional"[tiab] OR "Cohort Studies"[MeSH] OR "longitudinal"[tiab] OR "prospective cohort"[tiab] OR "follow-up stud*"[tiab] OR "repeated survey*"[tiab] OR "repeat cross-section*"[tiab])

AND

("2 year*"[tiab] OR "two year*"[tiab] OR "24 month*"[tiab] OR "3 year*"[tiab] OR "four year*"[tiab] OR "5 year*"[tiab] OR "long-term follow*"[tiab]))

**References**

1· Harris PA, et al· The REDCap consortium: Building an international community of software platform partners· *J Biomed Inform*· 2019;95:103208·

2· Harris PA, et al· Research electronic data capture (REDCap)--a metadata-driven methodology and workflow process for providing translational research informatics support· *J Biomed Inform*· 2009;42(2):377–381·

3· R Core Team· R: A Language and Environment for Statistical Computing· 2024· https://www·R-project·org/·

4· Hijmans RJ, et al· geosphere: Spherical Trigonometry· 2024· https://CRAN·R-project·org/package=geosphere· Accessed November 24, 2025·

5· Pebesma E, et al· sp: Classes and Methods for Spatial Data· 2025· https://cran·r-project·org/web/packages/sp/index·html· Accessed November 24, 2025·

6· Hijmans RJ, et al· raster: Geographic Data Analysis and Modeling· 2025· https://CRAN·R-project·org/package=raster· Accessed November 24, 2025·

7· Pebesma E, Bivand R· *Spatial Data Science: With Applications in R*· New York: Chapman and Hall/CRC; 2023·

8· Wickham H, et al· tidyr: Tidy Messy Data· 2024· https://CRAN·R-project·org/package=tidyr· Accessed November 24, 2025·

9· Wickham H, et al· dplyr: A Grammar of Data Manipulation· 2023· https://CRAN·R-project·org/package=dplyr· Accessed November 24, 2025·

10· Grolemund G, Wickham H· Dates and Times Made Easy with lubridate· *J Stat Softw*· 2011;40(3):1–25·

11· Wickham H, et al· scales: Scale Functions for Visualization· 2023· https://CRAN·R-project·org/package=scales>· Accessed November 24, 2025·

12· Wickham H, et al· readr: Read Rectangular Text Data· 2024· https://CRAN·R-project·org/package=readr· Accessed November 24, 2025·

13· Wickham H· *ggplot2: Elegant Graphics for Data Analysis*· Cham: Springer International Publishing; 2016·

14· Sievert C· *Interactive Web-Based Data Visualization with R, plotly, and shiny*· New York: Chapman and Hall/CRC; 2020·

15· Cheng J, et al· leaflet: Create Interactive Web Maps with the JavaScript “Leaflet” Library· 2024· https://CRAN·R-project·org/package=leaflet· Accessed November 24, 2025·

16 Bates D, Mächler M, Bolker B, Walker S. Fitting Linear Mixed-Effects Models Using lme4. Journal of Statistical Software 2015; 67: 1–48.

17 Bolker B, Robinson D. broom.mixed: Tidying Methods for Mixed Models. 2026 https://CRAN.R-project.org/package=broom.mixed.

18 Dorai-Raj S. binom: Binomial Confidence Intervals for Several Parameterizations. 2022 https://CRAN.R-project.org/package=binom.

19 Hartig F. DHARMa: Residual Diagnostics for Hierarchical (Multi-Level / Mixed) Regression Models. 2024 https://CRAN.R-project.org/package=DHARMa.

20 Lüdecke D. sjPlot: Data Visualization for Statistics in Social Science. 2025 https://CRAN.R-project.org/package=sjPlot.

21 Sjoberg DD, Whiting K, Curry M, Lavery JA, Larmarange J. Reproducible Summary Tables with the gtsummary Package. The R Journal 2021; 13: 570–80.

22· Global Administrative Areas (GADM)· https://gadm·org/· Accessed November 24, 2025·
