## Supplementary material for "Dynamics of *Plasmodium falciparum* asymptomatic asexual and sexual stages across transmission seasons in a rural and high-transmission setting in Burkina Faso: a two-year longitudinal cohort study with cross-sectional surveys": STROBE statement

| STROBE Statement—Checklist of items that should be included in reports of *cross-sectional studies* [1] | | | | |
| --- | --- | --- | --- | --- |
|  | Item No | Recommendation | Checked | |
| **Title and abstract** | 1 | (*a*) Indicate the study’s design with a commonly used term in the title or the abstract | | OK |
|  |  | (*b*) Provide in the abstract an informative and balanced summary of what was done and what was found | | OK |
| Introduction | | | |  |
| Background/rationale | 2 | Explain the scientific background and rationale for the investigation being reported | | OK |
| Objectives | 3 | State specific objectives, including any prespecified hypotheses | | OK |
| Methods | | | |  |
| Study design | 4 | Present key elements of study design early in the paper | | OK |
| Setting | 5 | Describe the setting, locations, and relevant dates, including periods of recruitment, exposure, follow-up, and data collection | | OK |
| Participants | 6 | (*a*) Give the eligibility criteria, and the sources and methods of selection of participants | | OK |
| Variables | 7 | Clearly define all outcomes, exposures, predictors, potential confounders, and effect modifiers· Give diagnostic criteria, if applicable | | OK |
| Data sources/ measurement | 8* | For each variable of interest, give sources of data and details of methods of assessment (measurement)· Describe comparability of assessment methods if there is more than one group | | OK |
| Bias | 9 | Describe any efforts to address potential sources of bias | | OK |
| Study size | 10 | Explain how the study size was arrived at | | OK |
| Quantitative variables | 11 | Explain how quantitative variables were handled in the analyses· If applicable, describe which groupings were chosen and why | | NA |
| Statistical methods | 12 | (*a*) Describe all statistical methods, including those used to control for confounding | | OK |
|  |  | (*b*) Describe any methods used to examine subgroups and interactions | | OK |
|  |  | (*c*) Explain how missing data were addressed | | OK |
|  |  | (*d*) If applicable, describe analytical methods taking account of sampling strategy | | NA |
|  |  | (*e*) Describe any sensitivity analyses | | NA |
| Results | | | |  |
| Participants | 13* | (a) Report numbers of individuals at each stage of study—eg numbers potentially eligible, examined for eligibility, confirmed eligible, included in the study, completing follow-up, and analysed | | OK |
|  |  | (b) Give reasons for non-participation at each stage | | OK |
|  |  | (c) Consider use of a flow diagram | | OK |
| Descriptive data | 14* | (a) Give characteristics of study participants (eg demographic, clinical, social) and information on exposures and potential confounders | | OK |
|  |  | (b) Indicate number of participants with missing data for each variable of interest | | OK |
| Outcome data | 15* | Report numbers of outcome events or summary measures | | OK |
| Main results | 16 | (*a*) Give unadjusted estimates and, if applicable, confounder-adjusted estimates and their precision (eg, 95% confidence interval)· Make clear which confounders were adjusted for and why they were included | | OK |
|  |  | (*b*) Report category boundaries when continuous variables were categorized | | OK |
|  |  | (*c*) If relevant, consider translating estimates of relative risk into absolute risk for a meaningful time period | | OK |
| Other analyses | 17 | Report other analyses done—eg analyses of subgroups and interactions, and sensitivity analyses | | OK |
| Discussion | | | |  |
| Key results | 18 | Summarise key results with reference to study objectives | | OK |
| Limitations | 19 | Discuss limitations of the study, taking into account sources of potential bias or imprecision· Discuss both direction and magnitude of any potential bias | | OK |
| Interpretation | 20 | Give a cautious overall interpretation of results considering objectives, limitations, multiplicity of analyses, results from similar studies, and other relevant evidence | | OK |
| Generalisability | 21 | Discuss the generalisability (external validity) of the study results | | OK |
| Other information | | | |  |
| Funding | 22 | Give the source of funding and the role of the funders for the present study and, if applicable, for the original study on which the present article is based | | OK |

*Give information separately for exposed and unexposed groups·

1·von Elm E, et al· The Strengthening the Reporting of Observational Studies in Epidemiology (STROBE) statement: guidelines for reporting observational studies· *PLoS Med*· 2007;4(10):e296·
